# Increased Systemic HSP70B Levels in Spinal Muscular Atrophy Infants

**DOI:** 10.1101/2020.11.20.20235325

**Authors:** Eric J. Eichelberger, Christiano R. R. Alves, Ren Zhang, Marco Petrillo, Patrick Cullen, Wildon Farwell, Jessica A Hurt, John F. Staropoli, Kathryn J. Swoboda

**Affiliations:** Department of Neurology, Center for Genomic Medicine, Massachusetts General Hospital, Boston, MA; Biogen, Cambridge, MA; Vertex Pharmaceuticals, Boston, MA

**Author notes:** Correspondence should be addressed to Kathryn J. Swoboda, Center for Genomic Medicine, Massachusetts General Hospital, Simches Research Building, 185 Cambridge Street, Boston, MA 02114, US, / / Cell. **Author Contributions:** EJE, CRRA, and KJS conceived and designed the study. EJE, RZ, JAH, PC and JFS carried out RNAseq experiments and analysis. MP and WF performed and analyzed neurofilaments experiments. EJE and CRRA carried out other experiments. KJS provided laboratory support and supervised the experiments. EJE and CRRA performed data analysis and drafted the manuscript. All authors interpreted the data, participated in manuscript review, and approved the final manuscript.

**Keywords:** Neuromuscular, Genetic Diseases, Biomarkers, Neurofilaments, Neurology

## Abstract

Despite newly available treatments for spinal muscular atrophy (SMA), novel circulating biomarkers are still critically necessary to track SMA progression and therapeutic response. To identify potential biomarkers, we performed whole-blood RNA sequencing analysis in SMA type 1 subjects under 1 year old and age-matched healthy controls. Our analysis revealed the *Heat Shock Protein Family A Member 7 (HSPA7)* as a novel candidate biomarker to track SMA progression early in life. Changes in circulating HSPA7 protein levels were associated with changes in circulating neurofilaments levels in SMA newborns and infants. Future studies will determine whether HSPA7 levels respond to molecular therapies.

## Introduction

With the federal recommendation to screen all newborns in the United States for spinal muscular atrophy (SMA) [1] and the emergence of three distinct effective molecular therapies to treat SMA newborns and infants at risk, identifying novel biomarkers to detect and monitor evidence of disease activity early in life is critical. Infants in the first year of life undergo rapid growth and transformational maturational events during a time when relying on clinical signs, symptoms or even established outcome measures are of insufficient sensitivity to predict the emergence of adverse clinical outcomes, whether due to an alteration of the underlying disease phenotype or as a secondary consequence of the therapy itself.

SMA is caused by mutations in the survival motor neuron 1 (*SMN1*) gene and the number of copies of *SMN2*, a paralog gene to *SMN1*, inversely correlates with phenotypic severity and is the primary disease modifier in SMA. Infants with 1-2 *SMN2* copies have prenatal or early neonatal onset of acute denervation that can result in severe outcomes, while symptom onset in those with 3 or more *SMN2* copies is much more variable, ranging from infancy to adulthood [2-5]. In spite of having three recent FDA-approved treatments for SMA, the lack of circulating biomarkers to track SMA progression and therapeutic consequences continues to impair our ability to ensure the best outcomes for SMA newborns.

Cerebrospinal fluid and plasma/serum neurofilament (NF) levels are markers of neuronal damage and have been recently identified as a potential prognostic and treatment responsive biomarker in SMA and other neurodegenerative diseases [6-8]. To further identify novel systemic biomarkers to predict and track SMA severity across a range of phenotypes, all newly diagnosed SMA patients, their siblings and their families are offered enrollment in the SPOT SMA Longitudinal Pediatric Data Repository (LPDR) database and linked biorepository [9-11]. The SPOT SMA LPDR was launched in 2016 to extend the Project Cure SMA LPDR, with appropriate modifications for anticipatory follow-up for SMA newborns and affected and unaffected siblings (NCT02831296). Here, we performed RNA sequencing and differential expression analyses in samples from SMA type 1 subjects under 1 year old and age-matched healthy control subjects to identify novel SMA biomarkers early in life. Our analysis identified for the first time the *Heat Shock Protein Family A Member 7 (HSPA7)* as a novel candidate biomarker to track SMA progression in the first year of life, indicating that its circulating protein levels are associated with NF levels in SMA newborns and infants.

## Materials and Methods

### Study Approval and Subjects

Written informed consent and parental consent were obtained from all participants under Institutional Ethics Review Boards at the University of Utah (protocol 8751) and Massachusetts General Hospital (protocol 2016P000469). We queried the Project Cure SMA and SPOT SMA LPDRs housed within the Research Electronic Data Capture Web Application at the Newborn Screening Translational Research Network for all RNA samples available from SMA subjects under 2 years of age not receiving any SMN-targeted molecular or gene therapy at the time of sample collection. Specifically, any subject receiving treatment with valproic acid (VPA), phenylbutyrate (PBA), nusinersen, onasemnogene abeparvovec and risdaplam were therefore excluded. For initial RNA sequencing exploratory analysis, we chose samples meeting strict quality control standards from 5 SMA type 1 infants and 5 healthy age-matched healthy controls under 1 year old (Cohort 1). We further validated our HSPA7 findings using all 51 RNA samples from 22 SMA subjects in our database (Cohort 2) using the same strict cutoffs for RNA integrity and purity. Validation of a selected target (HSP70B protein encoded by *HSPA7* gene) protein levels were performed to determine a potential correlation with NF levels (Cohort 3). **Table 1** presents the distribution of sex, age, SMA type, and *SMN2* copies for these cohorts. *SMN1* and *SMN2* copy numbers were determined using quantitative polymerase chain reaction as previously described [12].

**Table 1.**
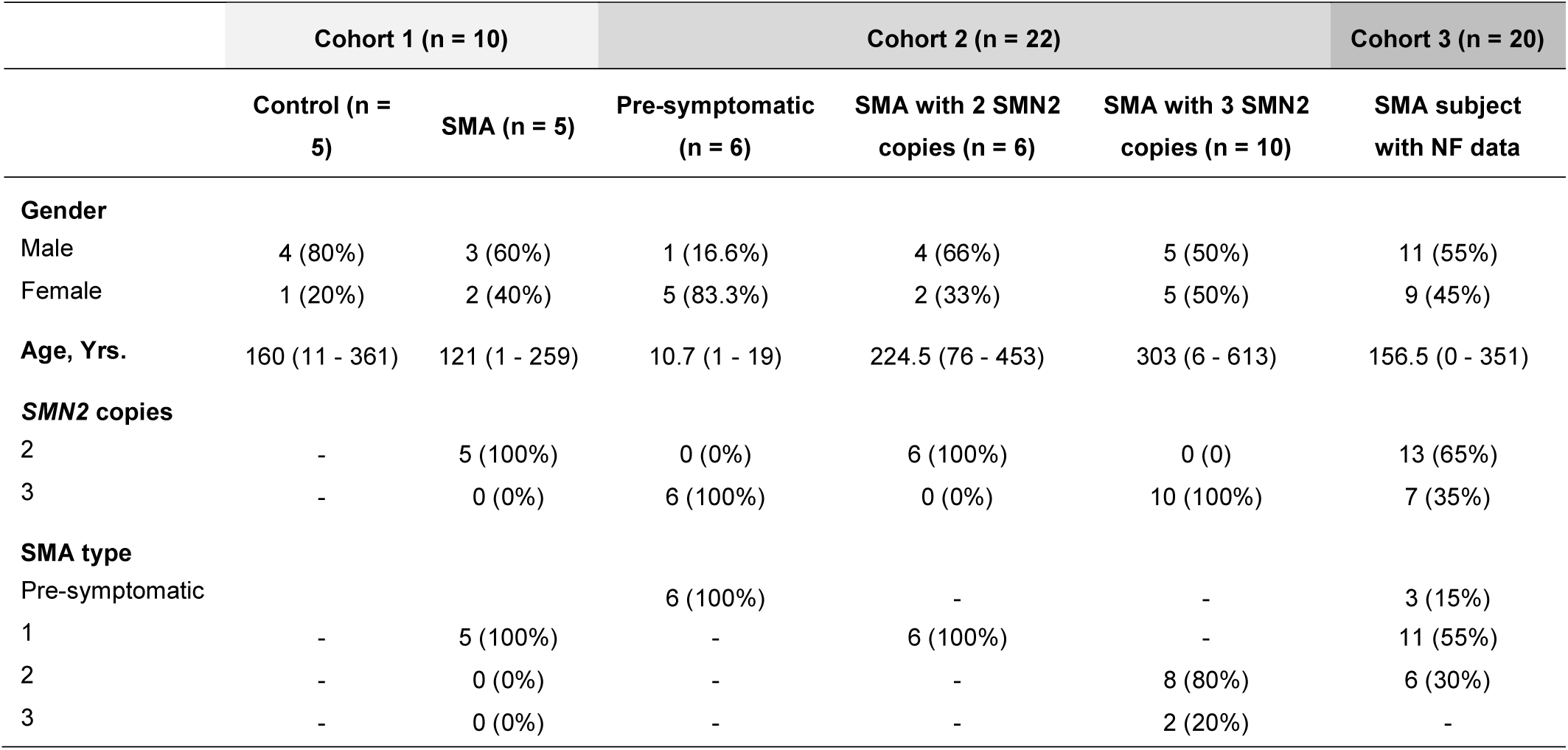
Characteristics of SMA and control subjects in each cohort studied.

### RNA Sequencing Analysis

RNA was extracted using the PAXgene Blood RNA kit (Qiagen, Hilden, Germany; 762164). RNA-sequencing libraries were prepared from 250 ng of DNase-treated total RNA using the TruSeq Stranded mRNA kit (Illumina). RNA sequencing FASTA files were trimmed and quality checked using FastQC. RNA-sequencing reads were then mapped to the hg38 human genome, which had been annotated with exons and introns. Counts for each gene were generated using the package STAR. A custom R script was created for differential expression analysis. Package DESeq2 was used for all differential expression at default parameters. Data were submitted to NCBI Gene Expression Omnibus (GEO) and will be released post-publication.

### Intron Retention Analysis

Read were aligned to genome version hg38 and annotation database Gencode V21 using the STAR algorithm (v. 2.4.0h). Resulting bam files were used to calculate the degree of intron retention per sample using the rMATS algorithm and custom intron annotation files, generated as described previously [13, 14]. Significantly retained introns were defined as FDR < 0.05 between SMA patients and controls.

Reverse-Transcription and Quantitative PCR of Sample mRNA. RNA was reverse transcribed using the High-Capacity cDNA Reverse Transcription Kit (Applied Biosystems, 4368814). cDNA was amplified using Power up SYBR green master mix (Applied Biosystems; A25742) in a QuantStudio 3 Real-Time PCR system (Applied Biosystems, A28567). The HPRT expression was used as housekeeping in all qPCR calculations. Primer sequences are as follow: HSPA7 Forward: GGCTAACAAGATCACCAATGACA; HSPA7 Reverse: TCGGCTTCATGAACCATCCT; HPRT Forward: GAAAAGGACCCCACGAAGTGT; HPRT Reverse: AGTCAAGGGCATATCCTACAA.

### HSP70B and Neurofilaments Level

Serum phosphorylated neurofilament heavy chain (pNf-H) concentrations were measured using a pNF-H enzyme-linked lectin assay (ProteinSimple, CA, USA) according to the manufacturer’s instruction and as previously described [8]. HSP70B levels were determined using a commercially available enzyme-linked immunosorbent assay (ELISA) kit (MyBioSource, CA, USA; MBS287601) according to manufacturer’s instructions.

### Statistical analysis

Data are presented as box plots (min to max) with dots as individual values. Statistical analyses were performed using R. Unpaired Student’s t tests or Kruskal–Wallis test were used to compare groups. Kendall rank correlation coefficient was used to evaluate correlations. Statistical significance was defined as p < 0.05.

## Results and Discussion

We analyzed a cohort of SMA patients with 2 *SMN2* copies with clinical symptoms early in life, which represented the natural history of the disease since subjects were not receiving any molecular or gene therapies at the sampling time. Whole blood RNA sequencing confirmed no expression of *SMN1* in all SMA patients (**Figure 1A**), with no significant difference in the *SMN2* expression between SMA subjects and controls (**Figure 1A**). We took advantage of this dataset to explore novel candidate biomarkers of SMA progression early in life. Differential expression analysis of exons showed 206 genes with p < 0.05 in SMA subjects as compared to controls, and these genes clustered together in healthy controls and 4 out of 5 SMA patients (**Figure 1B**). Using a more restricted cutoff of false discover rate (FDR) at 0.2 (excluding *SMN1*), we found 5 genes upregulated in SMA subjects as compared to controls (**Figure 1C**). Among these genes, *HSPA7* was the only gene expressed in whole blood in all subjects and consistently upregulated in all SMA patients (**Figure 1C**). RT-qPCR analysis validated these findings, showing a 2.7-fold increase in *HSPA7* mRNA levels in SMA subjects as compared to controls (**Figure 1D**).

**Figure 1.**
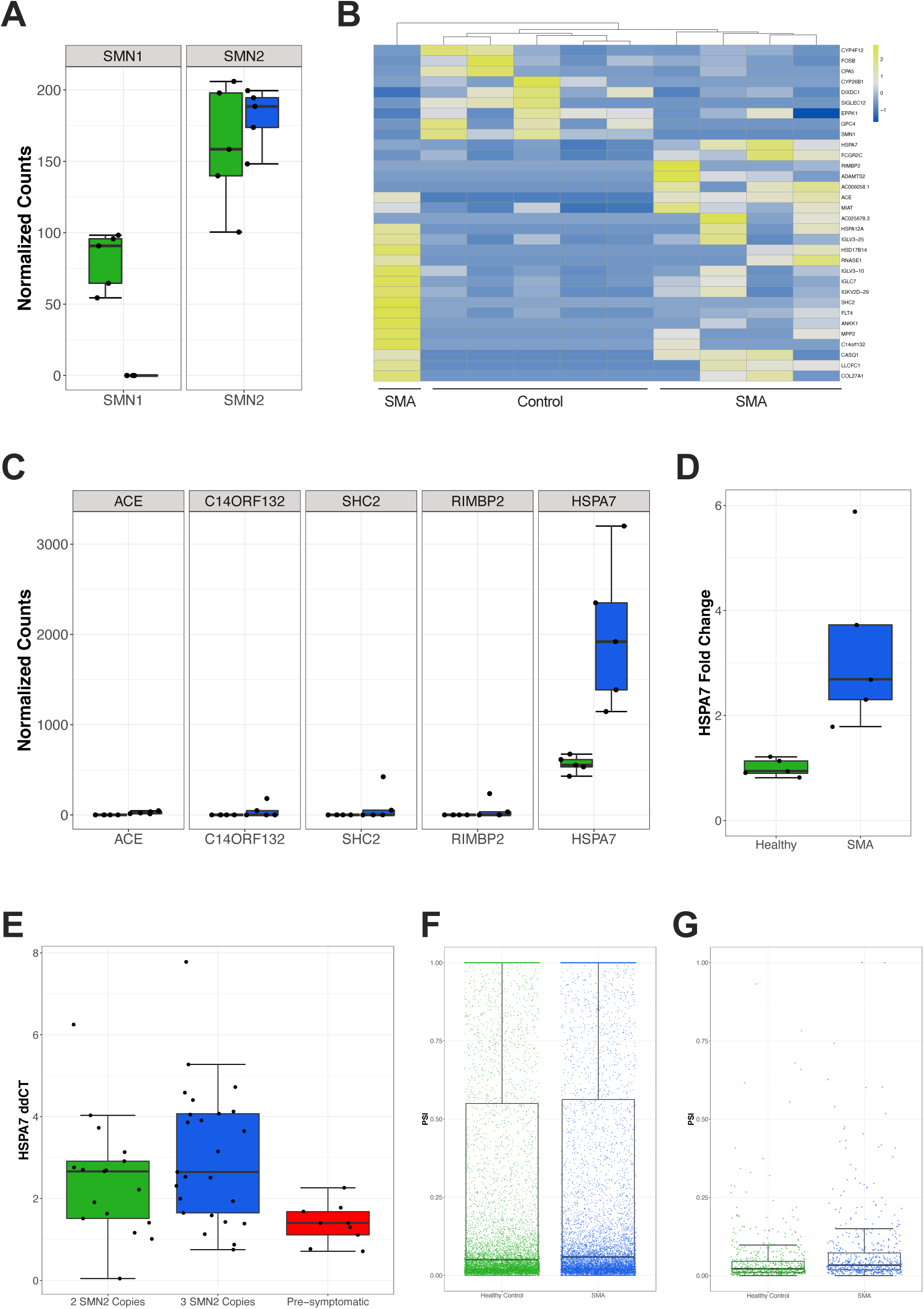
RNA sequencing reveals increased HSP70B mRNA levels in the whole blood of SMA Infants. **A:** Negative binomial normalized counts of *SMN1* and *SMN2* genes in SMA versus healthy controls. **B:** Heatmap plot of normalized counts for genes with p < 0.01 and an absolute value of log2 fold change > 1.2. **C:** Normalized counts of SMA and healthy patients for differentially expressed genes with FDR < 0.2. **D:** Quantitative RT-qPCR for HSPA7 mRNA levels. p = 0.02. **E:** HSPA7 mRNA levels in a cohort of SMA patients divided in symptomatic SMA subjects with only 2 *SMN2* copies, symptomatic SMA subjects with 3 *SMN2* copies, and pre-symptomatic SMA subjects with 3 *SMN2* copies. p = 0.01 and p = 0.003 when comparing pre-symptomatic SMA subjects with symptomatic SMA subjects with only 2 SMN2 copies and symptomatic SMA subjects with 3 *SMN2* copies, respectively. **F and G:** Comparison between SMA and controls for total (**F;** p < 0.001) and U12 (**G;** p < 0.001) intron retention in the whole blood. Individual values indicate each intron retained in individual samples. PSI: percentage spliced-in values.

We sought to evaluate the *HSPA7* mRNA expression in an independent cohort of SMA subjects with whole-blood RNA samples collected in the two years of life. We quantified *HSPA7* expression in a total of 51 samples from 22 different subjects and next divided samples into three groups: 1) symptomatic SMA subjects with 2 *SMN2* copies, 2) symptomatic SMA subjects with 3 *SMN2* copies, and 3) pre-symptomatic SMA subjects with 3 *SMN2* copies (**Table 1**). Symptomatic SMA subjects with 2 or 3 *SMN2* copies had significant higher *HSPA7* mRNA expression than pre-symptomatic SMA subjects (**Figure 1E**). There was no significant difference between symptomatic subjects with 2 and 3 SMN2 copies (**Figure 1E**). Based on these findings, we raised the hypothesis that *HSPA7* expression could be circulating biomarker for SMA.

In an additional analysis, we determined whether changes in SMA subjects would be associated to increased intron retention in whole blood. SMN plays a critical role in the assembly of small nuclear ribonucleoproteins (snRNPs), controlling correct spliceosomal assembly Increased intron retention occurs in murine models of SMA and in human cellular models with depleted of SMN [13, 15]. However, there are no studies demonstrating intron retention in systemic tissues in SMA subjects, including the whole blood which could be useful in the clinical setting to track disease progression. Our current analysis revealed that SMA infants display retained introns in whole blood when compared to controls (**Figure 1F**) including main differences in the atypical class of spliceosomal introns U12-type (**Figure 1G**).

*HSPA7* is a single exon gene expected to encode the heat shock 70kDa protein 7 (HSP70B; UniProtKB accession number P48741). There is evidence showing that *HSPA7* is a functional gene and the *HSPA7* mRNA can be transcribed, but little is known about the potential function (if any) of the HSP70B protein [16-18]. Here, we used a commercially available ELISA to measure the serum HSP70B protein levels in SMA subjects under 1 year old. These experiments revealed that serum HSP70B protein is detectable at concentrations ranging from 6.9 to 26.2 ng/mL in SMA newborns and infants. We therefore aimed to determine a potential correlation between serum HSP70B protein levels and neurofilaments, as circulating neurofilaments are markers of neuronal damage and have been established as a prognostic and treatment responsive biomarker in SMA [6-8]. We analyzed 37 serum samples available in our database with available pNf-H data in SMA subjects under 1 year old not receiving therapies. We observed a positive correlation between serum pNf-H and HSP70B protein levels (**Figure 2A**). Because we also observed an extensive range in the pNf-H concentrations (0.98 to 29.06 ng/mL), we next divided this cohort of samples into two subgroups based on the median of the pNf-H data (**Figure 2B**). Remarkably, SMA patients with high serum pNf-H levels also displayed significantly higher HSP70B protein levels as compared to SMA patients with low pNf-H levels (**Figure 2C**).

**Figure 2.**
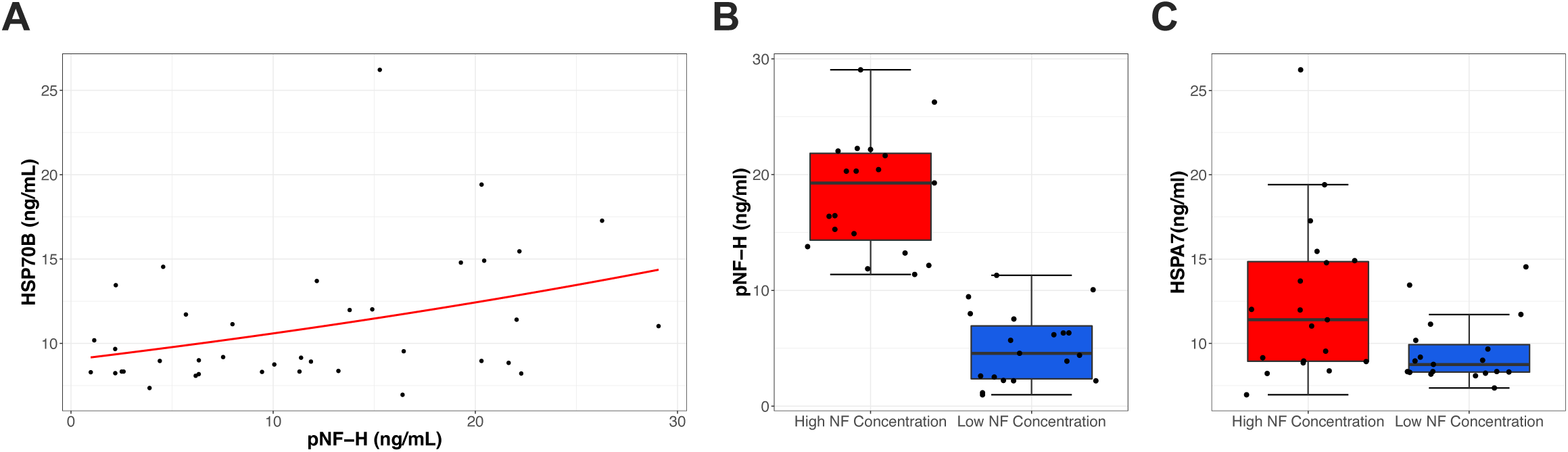
Association between HSPA7 protein levels and neurofilament concentrations. **A:** Association between serum HSP70B protein levels and neurofilament concentrations. Kendall rank correlation coefficient revealed p = 0.018. **B:** Division of samples in two groups based on low and high neurofilament concentrations in SMA patients. **C:** HSP70B protein levels in samples previously divided based on neurofilament levels. p = 0.01 between groups.

These novel findings demonstrate increased systemic HSP70B levels and intron retention in SMA newborns and infants. Because circulating HSP70B levels can be precisely measured using ELISA experiments, we will include these measurements in our routine analysis with SMA subjects to further acquire longitudinal follow-up data and determine the effects of molecular and/or gene therapies. We also encourage other additional studies to test circulating HSP70B levels in SMA patients, which can be in principle a new biomarker to track SMA progression early in life.

## Data Availability

RNA sequence data were submitted to NCBI Gene Expression Omnibus (GEO) and will be released post-publication.

## Acknowledgments

We are grateful to all the patients and families who participated in this study. We would like to thank Chao Sun and Will Chen for helpful discussions during this study. CRRA received a fellowship from the MGH Executive Committee on Research. KJS received financial support from NIH NICHD R01HD054599, NIH NINDS R21NS108015, Biogen and Cure SMA.

## Notes

**Conflict of Interest Statement:** EJE, CRRA and RZ report no conflict of interest. MP, WF, PC and JAH are employees of Biogen and hold stock/stock options in Biogen. JFS was employed at Biogen at the time the work was performed and holds stock/stock options in Biogen; he is now employed at Vertex Pharmaceuticals, Boston. KJS is on the scientific advisory board for Cure SMA and is a consultant for Biogen, Roche and AveXis. KJS receives clinical trial funding from AveXis and Biogen.

### Competing Interest Statement

EJE, CRRA and RZ report no conflict of interest. MP, WF, PC and JAH are employees of Biogen and hold stock/stock options in Biogen. JFS was employed at Biogen at the time the work was performed and holds stock/stock options in Biogen; he is now employed at Vertex Pharmaceuticals, Boston. KJS is on the scientific advisory board for Cure SMA and is a consultant for Biogen, Roche and AveXis. KJS receives clinical trial funding from AveXis and Biogen.

### Author Declarations

. Written informed consent and parental consent were obtained from all participants under Institutional Ethics Review Boards at the University of Utah (protocol 8751) and Massachusetts General Hospital (protocol 2016P000469).

